# Racial/Ethnic Disparities in Suicide Attempt Risk in New York City Female Youth

**DOI:** 10.1101/2025.06.02.25327628

**Authors:** Kara J. Emery, Md Mouinul Islam, Ezra M Solidum, John Dixon, Arielle H. Sheftall

## Abstract

**Objectives:** We investigated disparities in suicide attempt risk among Black/African American, Hispanic/Latinx, and White female youth in New York City (NYC) compared to National trends in the United States (US).

**Methods:** This study used data from the CDC Youth Risk Behavior Survey (YRBS) from 1997-2023. We analyzed self-reported racial/ethnic category, biological sex, and suicide attempt status. Suicide attempt risk and risk ratios were determined using logistic regression.

**Survey Population:** Our sample included 83,347 female youth Nationally (age in years M = 15.98, SD = 1.22) and 29,802 in NYC (age in years M = 15.56, SD = 1.24), with 9,480 (National) and 3,864 (NYC) of these females having attempted suicide at least once.

**Results:** Both samples showed the highest increases in suicide attempt risk among Black/African American female youth. While National and NYC samples showed significant racial disparities, these disparities were higher in NYC, especially during the COVID-19 pandemic.

**Conclusions:** This study showed pervasive racial disparities in suicide attempt risk for female youth and is one of the first to directly analyze local-level risk in female youth from a majority Black/African American and Hispanic/Latina sample.

## Introduction

In the United States (US), suicide is the second leading cause of death among youth aged 10 to 24 years of age.^1^ Over the past two decades, suicide rates in this age group have increased around 50%.^2^ Historically, youth suicide rates have been higher for males than females^2^, in part because the lethality of their attempts is higher.^3^ Recent trends reveal that this gap is narrowing and that suicide rates in females have been increasing faster than those of males^2–4^ across nearly all races and ethnicities.^5^ From 2011-2021, female youth consistently exhibited increasingly higher rates of suicidal ideation and suicide attempts compared to their male counterparts, which are both associated with suicide death.^6^ Addressing these increasing trends in female youth is critical, however, the current literature on how to do so is lacking.

Recent reports suggested potential underlying causes for the emerging sex disparities in suicide and suicidal behaviors. In 2023, the Centers for Disease Control and Prevention (CDC) released a report pointing to the rising mental health challenges among young females.^7^ Current data from the 2021 National CDC Youth Risk Behavior Survey (YRBS) showed that female teens faced the highest levels of sexual violence, sadness, and hopelessness ever reported, with persistent increases in the last decade.^3,8^

Another trend that has emerged in the field of youth suicidal behaviors is the presence of racial disparities.^9^ Although current research analyzing national trends indicated suicide rates are declining among White youth (ages 10-19),^10^ suicide rates in Black youth are rapidly increasing.^11^ Black females, in particular, have experienced the most rapid increase in suicide rates of any demographic group,^12^ with an increase of 182% over the last decade.^13^ Despite these trends, there are very few, if any, culturally tailored prevention efforts to address these alarming increases.^12^

National datasets analyzing youth suicidal thoughts and behaviors tend to have a much higher proportion of White youth in their samples than is representative of the populations in certain local communities. Trends in suicide rates conducted at the national level might be obscuring the impact on youth of color in these communities. For example, historic national trends in suicide have shown higher rates among White youth compared to their Black/African American and Hispanic/Latinx youth counterparts.^2–3^ While there have been recent efforts to calling attention growing racial disparities in STB, more localized and representative samples demonstrating the specific contextual needs of these populations are needed.^14–15^

With this in mind, the current study used data from the YRBS over the years 1997-2023 to determine whether suicide attempt risk is higher in Black/African American and Hispanic/Latinx compared to White adolescent females, and the extent of racial disparities in suicide attempt risk in NYC versus national samples. NYC provides a representative sample of these racial/ethnic groups, with almost half of its female youth residents identifying as either Black/African American or Hispanic/Latina.^16^ The boroughs of NYC are some of the most demographically diverse counties in the nation,^17^ with most having majority populations of color and together spanning a wide range of socioeconomic statuses (see Supplementary Table 1). Furthermore, even though NY is not among the states ranked highest in terms of overall suicide rates, the burden of suicidality in terms of total lives affected remains significantly high given its population density.^18^ This study takes an intersectional and selective approach to comparing suicide attempt risk in female youth which can provide insight on necessary targeted suicide prevention efforts.^19^

## Methodology

We followed the Strengthening the Reporting of Observational Studies in Epidemiology (STROBE) reporting guidelines for cross-sectional designs.^20^ We used publicly available data from the CDC, and in accordance with the Common Rule, Institutional Review Board (IRB) approval and informed consent were not necessary.

### Data Source and Participants

The CDC developed the Youth Risk Behavior Surveillance System (YRBSS) to monitor health behaviors and conditions among high school students throughout the US.^21^ The Youth Risk Behavior Survey (YRBS) questionnaire is a cross-sectional survey administered biennially during odd numbered years from 1991-2023 among students in grades 9-12 enrolled in U.S. public and private schools.^22^ Student participation in the survey is anonymous and voluntary through a self-administered, computer-scannable questionnaire.

National and site-level YRBS data are available in a combined data set online through the CDC. Sites used a two-stage cluster sampling design to produce a representative sample of students in their jurisdiction. The CDC provides sampling weights (based on student sex, race/ethnicity, and grade) to adjust for nonresponse and oversampling for each jurisdiction. Comprehensive reports of the study design and sampling procedures are available elsewhere.^22^

We analyzed data from the CDC’s YRBS National District Combined Dataset^23^ for national- and NYC-level trends. NYC site-level analyses included the following districts: Borough of Bronx, Borough of Brooklyn, Borough of Manhattan, Borough of Queens, and Borough of Staten Island. Our analytic sample included data from the years 1997-2023 for National and NYC-wide trends and 2003-2019 for site-level trends (given those were the only years available). We only included complete cases in our analyses without missing values for sex, race, or suicide attempt status. This included 83,347 (after 11.95% missing cases) female adolescents in the national sample and 35,383 (after 16.53% missing cases) in the NYC sample.

### Measures

YRBS race and ethnicity questions and their multiple choice responses have changed from 1991-2023. Therefore, the CDC created race/ethnicity variables in their combined dataset which standardized responses to these questions over time. Our dataset included the four-level race/ethnicity variable which provided the following categories: ‘White’, ‘Black or African American’, ‘Hispanic/Latino’, and ‘All Other Races’. There was only one question in the combined YRBS that assessed sex (“What is your sex?”), and none which assessed female gender identity. Our sample included all adolescents who responded as female for their sex and as ‘White’, ‘Black or African American’, or ‘Hispanic/Latino’ for race/ethnicity. We excluded the ‘All Other Races’ category in our analyses because it was consistently the smallest sample and was non-specific to which racial categories were represented.

One question in the YRBS assessed suicide attempt history: “During the past 12 months, how many times did you attempt suicide?”. We analyzed the CDC’s dichotomized version of this variable with “no” = “0 times” and “yes” = all other response options (i.e., one time, 2 or 3 times, 4 or 5 times, six or more times). This question was determined as a reliable measure by previous work.^24^

### Statistical Analysis

Logistic regression analyses were implemented to estimate linear and non-linear trends, model-adjusted risks, and risk ratios in suicide attempt risk. All analyses followed recommended procedures for trend and risk analysis with complex survey data.^25^ School grade was included as a covariate across all models to account for age- and cohort-related effects. To analyze linear and non-linear trends, we treated suicide attempt status as the dichotomous outcome variable and defined the survey year as a continuous variable with linear, quadratic, and cubic terms. Linear and non-linear terms were orthogonalized and tested sequentially. Segmented regression was used to automate identification of the year(s) when breakpoints occurred for non-linear risk trends. For this analysis, we used the *selgmented* function of the *segmented* package in R which identifies breakpoints according to the BIC criterion, or sequential hypothesis testing. Breakpoints were estimated based on model-adjusted risk estimates.

Model-adjusted risks and their 95% confidence intervals were estimated based on each logistic regression model’s predictive marginal means which accounted for the complex survey design using *svypredmeans* in R (cite). Risk ratios (RRs) were estimated from these model-adjusted risks, and their 95% confidence intervals were first calculated on the log scale and then exponentiated.^25^ RR analyses compared suicide attempt risk between White, Black/African American, and Hispanic/Latina females using White females as the reference group. RRs significantly greater than 1 indicated a higher suicide attempt risk compared to White females.

Odds ratios (ORs), risk ratios (RRs), and their corresponding 95% CIs are reported. Statistical significance was set to P < 0.05 with Wald 𝝌2 tests using design-adjusted coefficient variance-covariance matrices for all results. All reported analyses accounted for the complex sampling design and were conducted using the survey library in R. All regression models were estimated separately for each racial/ethnic group unless otherwise specified. See our linked code and Supplementary Material for additional details.

## Results

### Sociodemographic Characteristics

Sample sizes by race/ethnicity overall and for suicide attempts are shown in Table 1. For the National sample, of the 83,347 females (age in years *M* = 15.98, *SD* = 1.22), the majority were White (weighted sample percentages: 65% White; 21% Hispanic/Latina; and 14% Black/African American). For the NYC sample, of the 35,383 females (age in years *M* = 15.56, *SD* = 1.24), the majority were Hispanic/Latina and Black/African American (weighted sample percentages for NYC: 44% Hispanic/Latina; 37% Black/African American; and 19% White). The NYC sample varied in racial/ethnic composition by district, but all boroughs included a majority of Black/African American and Hispanic/Latina females, except for Staten Island.

**TABLE 1.** UNWEIGHTED SAMPLE SIZES AND WEIGHTED PERCENTAGES FOR FEMALE ADOLESCENTS ACROSS ALL GRADES BY RACE/ETHNICITY: US AND NYC YRBS, 1997–2023

|  | **All Groups, No. (%)** |  | **Black/African American, No. (%)** |  | **Hispanic/Latina, No. (%)** |  | **White, No. (%)** |  |
| --- | --- | --- | --- | --- | --- | --- | --- | --- |
|  | **Total<sup>a</sup>** | **Suicide Attempts<sup>b</sup>** | **Total** | **Suicide Attempts** | **Total** | **Suicide Attempts** | **Total** | **Suicide Attempts** |
| **US** | 83347 | 9480 (11) | 16973 (14) | 1883 (11) | 24277 (21) | 3481 (14) | 42097 (65) | 4116 (9) |
| **NYC** | 35383 | 3864 (11) | 10883 (37) | 1080 (10) | 18541 (44) | 2386 (13) | 5959 (19) | 398 (7) |
| **Bronx<sup>d</sup>** | 6546 | 765 (12) | 1833 (34) | 161 (9) | 4481 (62) | 585 (14) | 232 (4) | 19<sup>c</sup> (9) |
| **Brooklyn** | 6513 | 748 (10) | 3192 (55) | 353 (11) | 2518 (27) | 350 (12) | 803 (18) | 45 (5) |
| **Manhattan** | 7625 | 799 (12) | 2255 (33) | 197 (8) | 4694 (53) | 555 (13) | 676 (13) | 47 (8) |
| **Queens** | 4145 | 437 (10) | 1246 (34) | 117 (8) | 2258 (51) | 271 (13) | 641 (16) | 49 (7) |
| **Staten Island** | 4974 | 430 (8) | 688 (16) | 54 (8) | 1608 (25) | 214 (14) | 2678 (58) | 162 (6) |

<sup>a</sup>The data as the unweighted counts for the total sample included in the statistical analyses. Weighted percentages indicate the percent of the borough sample for each given racial group after accounting for the survey design.
<sup>b</sup>The data represents the unweighted counts for the total number of individuals in each sample who indicated at least one suicide attempt. Weighted percentages of attempters indicate the percent of attempters for the weighted sample of a given racial group after accounting for the survey design.
<sup>c</sup>Given the small sample size of White female adolescents in the Bronx, results for this borough should be interpreted with caution.

**TABLE 2.**
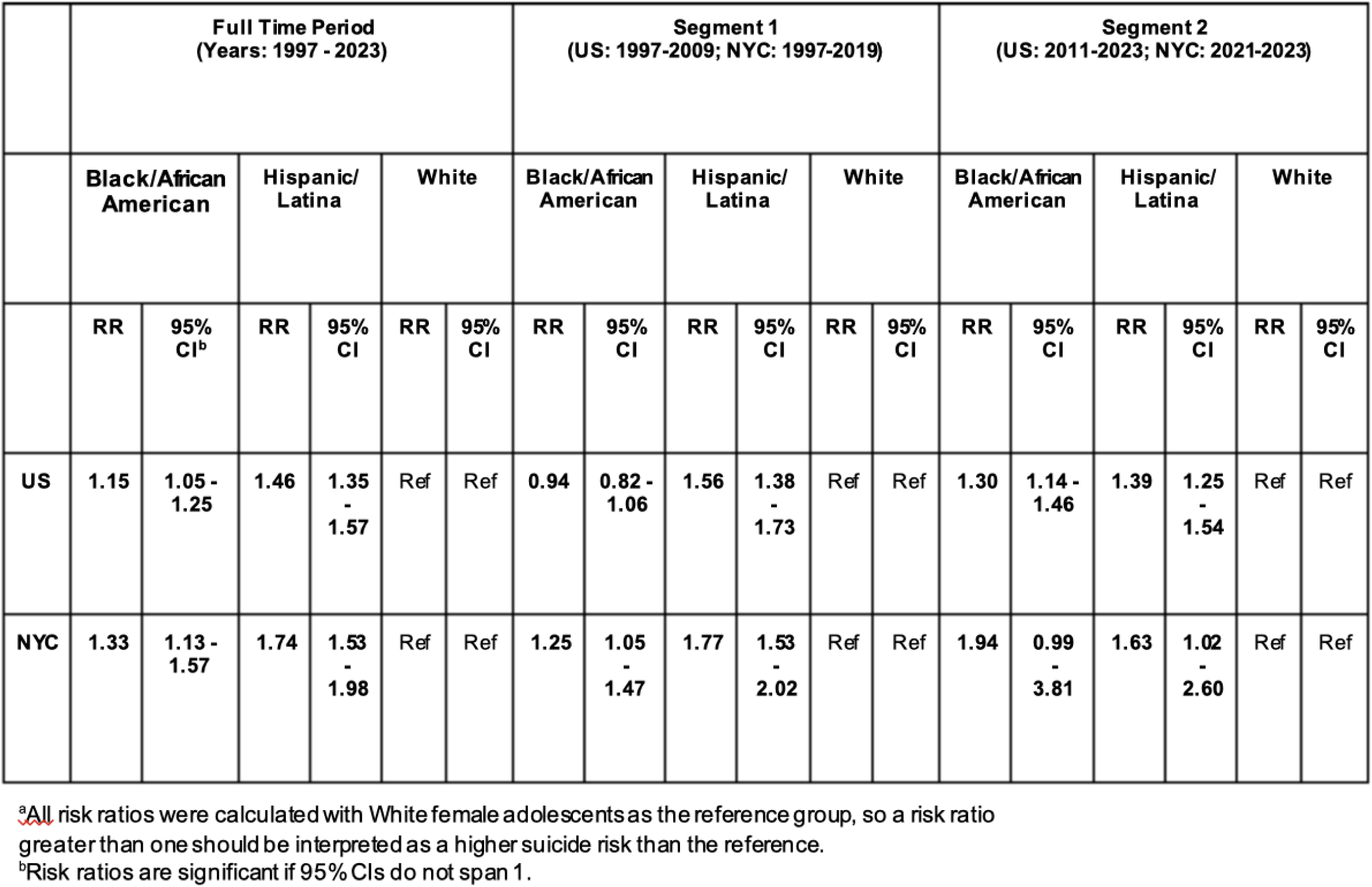
RISK RATIOS FOR SUICIDE ATTEMPTS AMONG BLACK/AFRICAN AMERICAN, HISPANIC/LATINA, AND WHITE FEMALE ADOLESCENTS: NATIONAL VS. NYC, 1997–2023 (REFERENCE GROUP: WHITE)^A^

From 1997-2023, there were 9,480 females who attempted suicide nationally and of those, 3,864 attempted suicide in NYC. Weighted suicide attempt percentages within each racial/ethnic group for the national sample included: 14.00% Hispanic/Latina; 10.90% Black/African American; and 9.29% White; and for the NYC sample: 13.21% Hispanic/Latina; 10.00% Black/African American; and 7.46% White.

### Suicide Attempt Risk

Figure 1 compared trends over time in suicide attempt risk between the US and NYC from 1997-2023 for Black/African American, Hispanic/Latina, and White female adolescents. Model-adjusted risks and ORs for all linear and nonlinear trends can be found in Supplementary Tables 2 and 3, respectively. Our evaluations confirm unique patterns in suicide attempt risk between National and NYC samples. For the US overall, significant increases in suicide attempt odds were found for Hispanic/Latina (quadratic: OR, 1.28; 95% CI, 1.02 to 1.61) and White female youth (quadratic: OR, 1.59; 95% CI, 1.29 to 1.96), with the highest increase for Black female youth (linear: OR, 2.26; 95% CI, 1.69 to 3.02). NYC showed only a trending increase in suicide attempt odds for Black female youth (linear: OR, 2.10; 95% CI, 1.00 to 4.43), and no significant changes for Hispanic/Latina or White female youth. There were significant racial disparities in suicide attempt odds nationally and in NYC. In the US overall from 1997 - 2023, Black/African American (RR, 1.15; 95% CI, 1.05 to 1.25) and Hispanic/Latina female youth (RR, 1.46; 95% CI, 1.35 to 1.57) showed significantly higher suicide attempt risk than White female youth (Reference group). Racial disparities were slightly higher in NYC compared to National estimates, with Black/African American (RR, 1.33; 95% CI, 1.13 to 1.57) and Hispanic/Latina female youth (RR, 1.74; 95% CI, 1.53 to 1.98) again showing significantly higher suicide attempt risk than White female youth.

**FIGURE 1.**
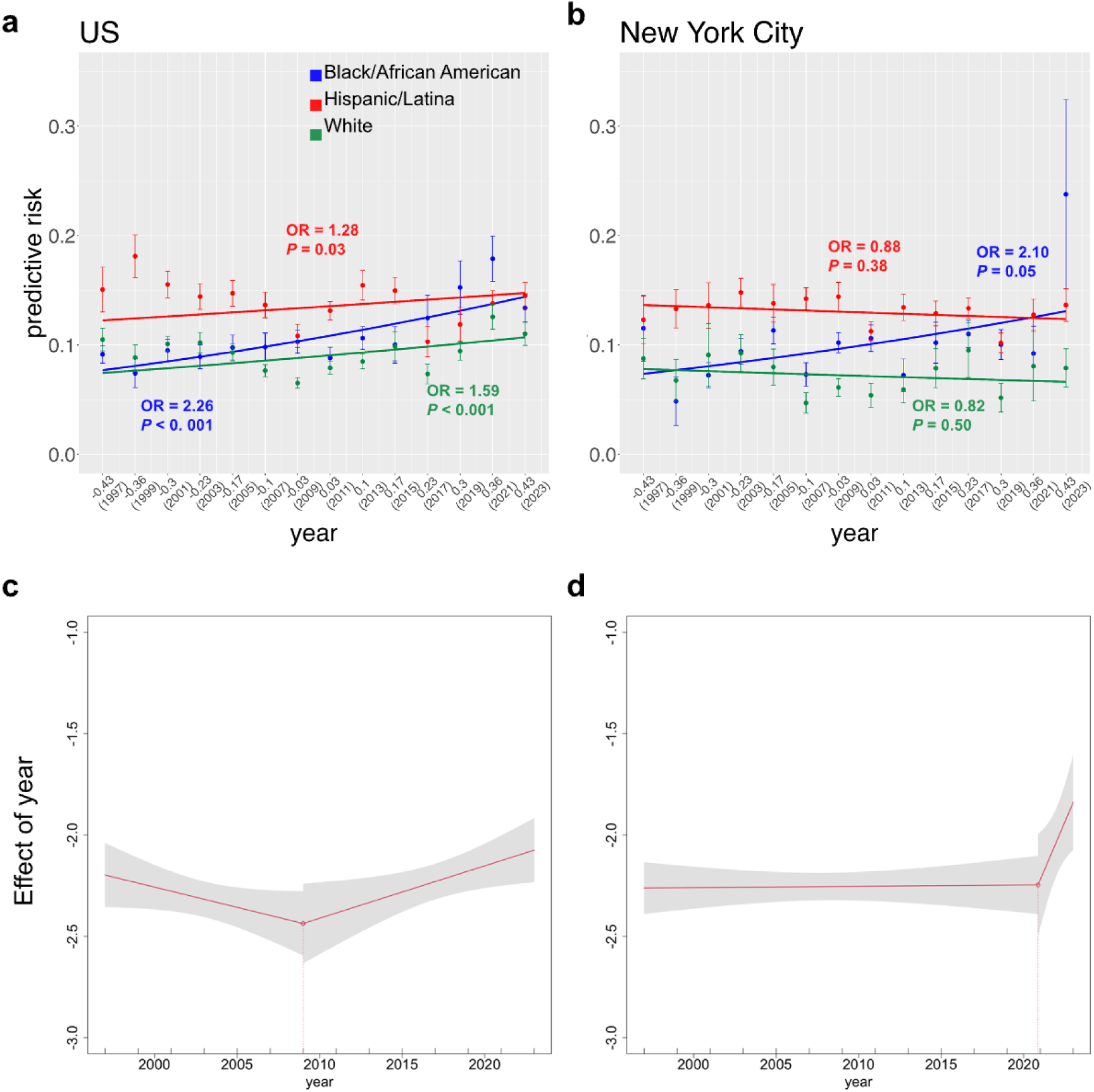
SUICIDE ATTEMPT RISK OVERTIME BY RACE/ETHNICITY AMONG FEMALE ADOLESCENTS: US VS. NYC, 1997–2023. **a-b**, Across all plots, colors corresponding to each racial/ethnic group are indicated in the legend. Points and their standard errors corresponded with the predicted suicide attempt risk for that given year by race/ethnicity (adjusted for grade). These predictive risks were estimated using the recommended method for complex survey designs^35^. The solid lines in each plot indicate the logistic regression results. These results correspond with the predicted risk by a given model evaluated at the linear contrast of each year. For trends which were **not** significant, lines were plotted accordingly to the best linear fit. For trends which were significant (linear and non-linear), lines correspond with the best fitting model’s predictive risk. See Supplementary Table 2 and 3 for all corresponding values. **c-d,** Segmented regressions were calculated across aggregated US and NYC samples to identify year(s) when breakpoints occurred. Breakpoints are indicated by the vertical dashed lines.

Racial disparities appeared to vary overtime (see Figure 1A-B), especially given the spike in suicide attempt risk for Black female youth in 2023 (model-adjusted risk, 23.79%; SE, 8.66%). Therefore, we aimed to determine whether there were significant changes in racial disparities in suicide attempt risk over the full time period. We applied segmented regression analyses to the US and NYC data separately (aggregating across all females within each group) to identify statistically significant breakpoints in suicide attempt risk (see Figure 1C-D). We found one significant breakpoint for the US at the year 2009 (SE, 2.48) and one significant breakpoint for NYC in the year 2020.87 (SE, 0.68). Notably the breakpoint for NYC corresponds with the beginning of the first school year during the COVID-19 pandemic. Nationally, RRs increased significantly for Black youth compared to White youth females from the first segment (years 1997-2009: RR, 0.94; 95% CI, 0.82 to 1.06) to the second (years 2011-2023: RR, 1.30; 95% CI, 1.14 to 1.46). There was a slight decrease between Hispanic/Latina and White females from the first segment (years 1997-2009: RR, 1.56; 95% CI, 1.38 to 1.73) to the second (years 2011-2023: RR, 1.39; 95% CI, 1.25 to 1.54) that did not reach significance. In NYC, RRs showed an increase for Black youth compared to White youth females from the first (years 1997-2019: RR, 1.25; 95% CI, 1.05 to 1.47) to the second segment (years 2021-2023: RR, 1.94; 95% CI, 0.99 to 3.81), but this increase did not reach significance, most likely due to the small number of time points in the second segment. Again, there was a slight decrease between Hispanic/Latina and White females from the first (years 1997-2019: RR, 1.77; 95% CI, 1.53 to 2.02) to the second segment (years 2021-2023: RR, 1.63; 95% CI, 1.02 to 2.60) that did not reach significance, also likely due to the small number of time points in the second segment.

To understand local trends in NYC, Figure 2 examined suicide attempt risk by borough from 2003-2019 in for Black/African American, Hispanic/Latina, and White female adolescents. Model-adjusted risks and ORs for all linear and non-linear trends can be found in Supplementary Tables 4 and 5, respectively. There were differences in suicide attempt risk trends by borough; more boroughs saw a significant decrease overtime in White female suicide attempt risk compared to the other racial/ethnic groups: Manhattan (cubic: OR, 0.22; 95% CI, 0.07 to 0.67), Queens (cubic: OR, 0.19; 95% CI, 0.07 to 0.51), and Staten Island (linear: OR, 0.57; 95% CI, 0.35 to 0.93). Hispanic/Latina females showed a significant decrease overtime in two boroughs: Brooklyn (quadratic: OR, 0.49; 95% CI, 0.29 to 0.83), and Manhattan (linear: OR, 0.47; 95% CI, 0.32 to 0.70). Black/African American females showed no significant linear or non-linear trends in suicide attempt risk over time in any borough. These results confirm that Black/African American and Hispanic/Latina female youth had higher rates of suicide attempts across most boroughs and were, in some cases, twice as likely to attempt suicide compared to their White counterparts.

**FIGURE 2.**
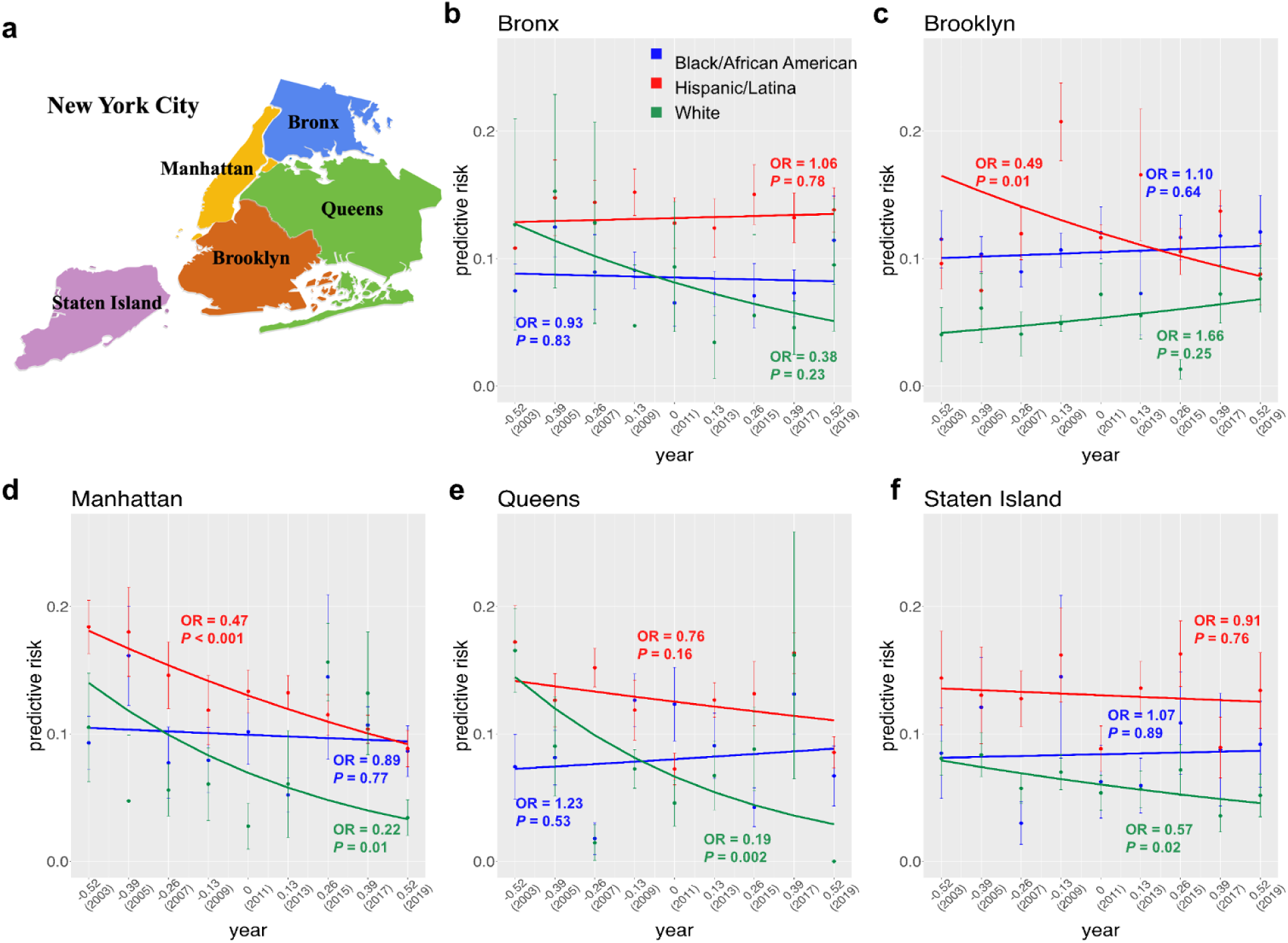
SUICIDE ATTEMPT RISK OVERTIME BY RACE/ETHNICITY AND NYC BOROUGH AMONG FEMALE ADOLESCENTS, 2003–2019. **a**, The location of each borough in New York City. **b-f,** Across all plots, colors corresponding to each racial/ethnic group are indicated in the legend. Points and their standard errors corresponded with the predicted suicide attempt risk for that given year by race/ethnicity (adjusted for grade). These predictive risks were estimated using the recommended method for complex survey designs^35^. The solid lines in each plot indicate the logistic regression results. These results correspond with the predicted risk by a given model evaluated at the linear contrast of each year. For trends which were **not** significant, lines were plotted accordingly to the best linear fit. For trends which were significant (linear and non-linear), lines correspond with the best fitting model’s predictive risk. See Supplementary Tables 4 and 5 for all corresponding values.

## Discussion

Results of this study offer a new perspective on the pervasive racial inequities present in suicide attempt risk among Black/African American, Hispanic/Latinx, and White female adolescents Nationally and in NYC. Historic national trends often de-emphasized suicide as an issue for youth of color and females, pointing to higher suicide mortality rates in White male youth.^2,3^ Historic national trends also de-emphasized suicide risk in New York where overall suicide mortality rates have been lower compared to other states.^18^ In this study, we confirmed significant increases in suicide attempt risk nationally for female youth, and that racial disparities are present, especially for Black youth females. We also confirmed that NYC provides a more diverse sample of female youth than is captured nationally, and that racial disparities in suicide attempt risk are higher in this group.

We learned that racial disparities in suicide attempt risk are further pronounced in NYC compared to National estimates, particularly for Black youth females. Black youth females were the only group in NYC to see a trending increase in suicide attempt risk, with a notable spike in predictive risk in 2023. Local trends by NYC borough also showed significant decreases for White and Hispanic/Latina female youth, while risk remained stagnant for Black youth. National suicide prevention strategies have historically focused on White youth^12^, which could have contributed to the relatively lower risk in this group. In NYC, given their relatively high STB risk, a targeted suicide prevention effort was established for Hispanic/Latina youth in NYC^26^, which could have contributed to the decreases witnessed in this group. The findings reported here demonstrate that suicide prevention, research, and intervention strategies that specifically target Black/African American females are vital to address these concerns and create sustainable change.

We also found a unique spike in suicide attempt risk for female youth in NYC during the COVID-19 pandemic, around the beginning of the school year (October 2020). This spike also marked a trending increase in racial disparities, particularly between Black/African American and White female adolescents. These findings align with a recent Centers for Disease Control and Prevention (CDC) report that found significant increases in suicides among Black, Hispanic, and Native American/Alaska Native groups during the COVID-19 pandemic.^27^ However, our results demonstrate that this spike in suicide attempt risk during the pandemic was unique to NYC compared to National estimates. These findings drive the need for increased local efforts in NYC focused on understanding what is influencing these COVID-related increases and their racial disparities, and developing targeted intervention strategies accordingly.

Recent work on the effectiveness of suicide intervention and prevention strategies has focused on identifying the level at which these strategies tend to be the most impactful.^28^ In this framework, suicide preventions and interventions are classified into three levels: universal, selective, and indicated. Universal strategies are applied to entire populations, selective prevention strategies are targeted toward populations at risk, and indicated strategies focus on individuals currently displaying STBs. A recent review showed that a combination of selective and indicated suicide preventions/interventions tended to be the most effective.^28^ Our findings aligned with this result, showing the importance of disaggregated approaches and how they might inform selective prevention strategies. Although it is important for national suicide prevention strategies in the US to address inequities by subgroups such as race/ethnicity and sex, designing a universal solution may be futile.

The results of this study and others emphasize the importance of confronting the rising mental health challenges in female youth using an intersectional approach.

Intersectionality frameworks define mutually reinforcing, interdependent systems of marginalization, rather than considering those systems in isolation.^29^ For example, female youth are at a higher risk of experiencing violence and trauma (significant risk factors for suicidal thoughts and behaviors) compared to their male counterparts.^8^ Youth of color are at an even higher risk of trauma exposure, especially for those in neighborhoods with higher levels of poverty.^30^ In both Black/African American and Latinx youth, racial discrimination is associated with higher suicidality, and this relationship is stronger in females versus males.^31–32^ The intersectionality of race/ethnicity, biological sex, and gender identity need to be fully examined to establish effective prevention efforts informed by the psychosocial and sociocultural factors associated with suicidal behaviors.^14^

This study has several limitations. One important limitation is that the YRBS sampling design included only youth attending public and private schools rather than all adolescents in this age group (e.g., homeschooled). Second, the suicide attempt measure is self-reported which leaves some potential for under- or over-reporting bias. Third, the NYC YRBS district-level data analyzed here is limited to 2019, which does not align with the National- and NYC-wide samples. Fourth, we relied on a simplified set of race/ethnicity categories for our analyses. Black/African American and Hispanic/Latina youth are diverse groups, and the current data did not offer a lens into how the specified trends might differ within these subgroups (e.g., Argentine descent). We also did not account for gender identity/expression or sexual orientation in these analyses, and it has been shown that LGBTQ+ youth have the highest rates of STB.^33^ Lastly, we only examined trends specific to NYC rather than including additional locations. However, our goal was to demonstrate the importance of examining local data for developing more targeted insights.

### Public Health Implications and Next Steps

This study is one of the first to directly compare national, city, and district-level suicide attempt risk disparities in female youth including a majority Black/African American and Hispanic/Latina sample. More research tailored to the discovery of specific risk factors and selective prevention strategies for Black/African American and Hispanic/Latina female youth to address these high rates of suicidal behaviors is needed, and approaching the problem from a disaggregated, local-level may be the best step forward. For example, Life is Precious, a Latina Suicide Prevention Program, with Locations in Brooklyn, The Bronx, Queens, and Washington Heights in New York City was founded in 2008 to address rising rates of STB in Latina youth. Our results show significant decreases in suicide attempt risk for Hispanic/Latina youth, suggesting these types of programs are worthwhile. Given the rising suicide attempt risk for Black youth females in NYC reported here, we suggest the development of a similar program specifically targeting STB in Black youth.

While recent work points to similar patterns of racial/ethnic disparities in Chicago^34^, future research should determine if our findings are replicable and relevant to other urban areas and other areas with majority populations of female youth of color.

Subsequent approaches should also leverage longitudinal datasets to better understand the causes of suicide attempt risk among these populations. Overall, determining the modifiable risks and protective factors for female youth of color at a community-level is necessary to adequately decrease rates of suicidal behaviors.

## Data Availability

All data produced are available online at https://www.cdc.gov/yrbs/data/index.html

