## Supplementary Material for "Racial/Ethnic Disparities in Suicide Attempt Risk in New York City Female Youth"

### Supplementary Table 1. Borough Census Profiles 2023

| | Race/Ethnicity | Female Population Aged 10-19 | Percentage of Females Aged 10-19 (%) | Median Household Income | Persons in Poverty (%) | Persons without Health Care Coverage (%) |
|---|---|---|---|---|---|---|
| NYC | | | | $76,577 | 18.20% | 5.80% |
| | All | 721,340 | | | | |
| | White | 113,726 | 15.77% | | | |
| | Black or African American | 106,540 | 14.77% | | | |
| | Hispanic or Latino | 279,936 | 38.81% | | | |
| | Asian | 59,670 | 8.27% | | | |
| | American Indian and Alaska Native | 5,532 | 0.77% | | | |
| | Native Hawaiian and Other Pacific Islander | 433 | 0.06% | | | |
| | Some Other Race Alone | 88,808 | 12.31% | | | |
| | Two or More Races | 66,696 | 9.25% | | | |
| | Race/Ethnicity | Female Population Aged 10-19 | Percentage of Females Aged 10-19 (%) | Median Household Income | Persons in Poverty (%) | Persons without Health Care Coverage (%) |
| Bronx | | | | $46,838 | 27.70% | 6.70% |
| | All | 150,001 | | | | |
| | White | 9,991 | 6.66% | | | |
| | Black or African American | 31,821 | 21.21% | | | |
| | Hispanic or Latino | 56,153 | 37.44% | | | |
| | Asian | 3202 | 2.13% | | | |
| | American Indian and Alaska Native | 1947 | 1.30% | | | |
| | Native Hawaiian and Other Pacific Islander | 252 | 0.17% | | | |
| | Some Other Race | 34,782 | 23.18% | | | |
| | Two or More Races | 11,853 | 7.90% | | | |

| | Race/Ethnicity | Female Population Aged 10-19 | Percentage of Females Aged 10-19 (%) | Median Household Income | Persons in Poverty (%) | Persons without Health Care Coverage (%) |
|---|---|---|---|---|---|---|
| Brooklyn | | | | $76,912 | 19.00% | 5.00% |
| | All | 181,198 | | | | |
| | White | 50,963 | 28.13% | | | |
| | Black or African American | 38,964 | 21.50% | | | |
| | Hispanic or Latino | 34,510 | 19.05% | | | |
| | Asian | 15,684 | 8.66% | | | |
| | American Indian and Alaska Native | 1,444 | 0.80% | | | |
| | Native Hawaiian and Other Pacific Islander | 19 | 0.01% | | | |
| | Some Other Race | 19,942 | 11.01% | | | |
| | Two or More Races | 19,672 | 10.86% | | | |
| | Race/Ethnicity | Female Population Aged 10-19 | Percentage of Females Aged 10-19 (%) | Median Household Income | Persons in Poverty (%) | Persons without Health Care Coverage (%) |
| Manhattan | | | | $101,078 | 16.50% | 4.00% |
| | All | 87, 788 | | | | |
| | White | 28,317 | 32.26% | | | |
| | Black or African American | 8,884 | 10.12% | | | |
| | Hispanic or Latino | 20,247 | 23.06% | | | |
| | Asian | 8,295 | 9.45% | | | |
| | American Indian and Alaska Native | 783 | 0.89% | | | |
| | Native Hawaiian and Other Pacific Islander | 67 | 0.07% | | | |
| | Some Other Race | 9,680 | 11.03% | | | |
| | Two or More Races | 11,515 | 13.12% | | | |

| | Race/Ethnicity | Female Population Aged 10-19 | Percentage of Females Aged 10-19 (%) | Median Household Income | Persons in Poverty (%) | Persons without Health Care Coverage (%) |
|---|---|---|---|---|---|---|
| Queens | | | | $81,929 | 13.70% | 7.80% |
| | All | 155,674 | | | | |
| | White | 24,669 | 15.85% | | | |
| | Black or African American | 22,503 | 14.46% | | | |
| | Hispanic or Latino | 39,962 | 25.67% | | | |
| | Asian | 27,132 | 17.43% | | | |
| | American Indian and Alaska Native | 1,376 | 0.88% | | | |
| | Native Hawaiian and Other Pacific Islander | 84 | 0.05% | | | |
| | Some Other Race | 22,658 | 14.55% | | | |
| | Two or More Races | 17,290 | 11.11% | | | |
| | Race/Ethnicity | Female Population Aged 10-19 | Percentage of Females Aged 10-19 (%) | Median Household Income | Persons in Poverty (%) | Persons without Health Care Coverage (%) |
| Staten Island | | | | $95,543 | 13.20% | 4.00% |
| | All | 38,403 | | | | |
| | White | 14,203 | 36.98% | | | |
| | Black or African American | 4,368 | 11.37% | | | |
| | Hispanic or Latino | 8,043 | 20.94% | | | |
| | Asian | 4,909 | 12.78% | | | |
| | American Indian and Alaska Native | 434 | 1.13% | | | |
| | Native Hawaiian and Other Pacific Islander | 11 | 0.03% | | | |
| | Some Other Race | 1,746 | 4.55% | | | |
| | Two or More Races | 4,689 | 12.21% | | | |

**Supplementary Table 2**. US and NYC Model-Adjusted Suicide Attempt Risk Estimates and Standard Errors (as Percentages) by Race/Ethnicity Among Female Adolescents, 1997–2023

| | | 1997 | 1999 | 2001 | 2003 | 2005 | 2007 | 2009 | 2011 | 2013 | 2015 | 2017 | 2019 | 2021 | 2023 |
|---|---|---|---|---|---|---|---|---|---|---|---|---|---|---|---|
| Black | US | 9.15 (0.81) | 7.42 (1.32) | 9.50 (1.00) | 8.93 (1.08) | 9.77 (1.14) | 9.81 (1.28) | 10.31 (1.05) | 8.83 (0.98) | 10.64 (1.05) | 10.02 (1.86) | 12.48 (2.09) | 15.26 (2.44) | 17.89 (2.08) | 13.39 (1.29) |
| | NYC | 11.53 (3.01) | 4.86 (2.21) | 7.24 (1.15) | 9.43 (1.18) | 11.33 (1.24) | 7.31 (1.07) | 10.21 (0.92) | 10.62 (1.22) | 7.24 (1.49) | 10.21 (1.87) | 11.01 (1.87) | 10.04 (1.37) | 9.23 (2.49) | 23.78 (8.66) |
| Latina | US | 15.07 (2.05) | 18.12 (1.95) | 15.54 (1.22) | 14.43 (1.17) | 14.73 (1.18) | 13.66 (1.17) | 10.83 (1.06) | 13.14 (0.84) | 15.46 (1.35) | 14.97 | 10.30 | 11.88 | 13.80 | 14.52 |

| | | | | | | | | | | | | | | | |
|---|---|---|---|---|---|---|---|---|---|---|---|---|---|---|---|
| | | | | | | | | | | (1.21) | (1.38) | (1.55) | (1.17) | (1.20) |
| | NYC | 12.30 (2.17) | 13.30 (1.74) | 13.62 (2.08) | 14.80 (1.30) | 13.81 (1.72) | 14.24 (1.00) | 14.42 (1.32) | 11.26 (0.86) | 13.44 (1.20) | 12.88 (1.13) | 13.36 (0.93) | 10.20 (0.89) | 12.76 (1.42) | 13.65 (1.49) |
| White | US | 10.50 (1.03) | 8.87 (1.13) | 10.10 (0.70) | 10.21 (0.93) | 9.31 (0.77) | 7.66 (0.56) | 6.53 (0.49) | 7.92 (0.57) | 8.49 (10.67) | 9.88 (1.33) | 7.35 (0.91) | 9.45 (0.83) | 12.57 (1.13) | 11.02 (1.05) |
| | NYC | 8.77 (1.85) | 6.77 (1.91) | 9.09 (2.86) | 9.27 (1.67) | 7.99 (1.65) | 4.70 (0.94) | 6.13 (0.81) | 5.40 (1.09) | 5.92 (1.21) | 7.88 (1.79) | 9.52 (2.49) | 5.18 (1.32) | 8.06 (3.18) | 7.90 (1.76) |

**Supplementary Table 3**. US and NYC Suicide Attempt Odds Ratios by Race/Ethnicity Among Female Adolescents, 1997–2023

| | | **Black/African American** | | | **Hispanic/Latina** | | | **White** | | |
|---|---|---|---|---|---|---|---|---|---|---|
| | | **OR** | **95% CI** | ***P*** | **OR** | **95% CI** | ***P*** | **OR** | **95% CI** | ***P*** |
| **US** | **L** | **2.26** | **1.69 – 3.02** | **<0.001** | 0.83 | 0.65 – 1.05 | 0.12 | 1.11 | 0.88 - 1.41 | 0.39 |
| | **Q** | 1.22 | 0.95 - 1.56 | 0.31 | **1.28** | **1.02-1.61** | **0.03** | **1.59** | **1.29 - 1.96** | **<0.001** |
| | **C** | 0.93 | 0.71 - 1.22 | 0.60 | 1.08 | 0.86 – 1.37 | 0.50 | 1.10 | 0.89 – 1.37 | 0.37 |
| **NYC** | **L** | 2.10 | 1.00 – 4.43 | 0.05 | 0.88 | 0.65 - 1.18 | 0.38 | 0.82 | 0.45 - 1.48 | 0.50 |
| | **Q** | 1.72 | 0.81 – 3.64 | 0.16 | 0.95 | 0.72 - 1.26 | 0.72 | 1.36 | 0.80 – 2.33 | 0.23 |
| | **C** | 1.50 | 0.71 - 3.17 | 0.29 | 1.24 | 0.95 - 1.64 | 0.12 | 1.05 | 0.58 – 1.89 | 0.87 |

**Supplementary Table 4**. Model-Adjusted Suicide Attempt Risk Estimates and Standard Errors (as Percentages) by NYC Borough and Race/Ethnicity Among Female Adolescents, 2003–2019

| | | 2003 | 2005 | 2007 | 2009 | 2011 | 2013 | 2015 | 2017 | 2019 |
|---|---|---|---|---|---|---|---|---|---|---|
| NYC | Black | 9.48 (1.31) | 11.29 (1.15) | 7.27 (1.08) | 10.13 (0.94) | 10.48 (1.10) | 7.15 (1.45) | 10.07 (1.86) | 10.91 (1.34) | 9.95 (1.38) |
| | Latina | 14.49 (1.54) | 13.76 (1.76) | 14.25 (1.00) | 14.51 (1.34) | 11.31 (0.94) | 13.46 (1.20) | 12.96 (1.11) | 13.40 (0.93) | 10.24 (0.90) |
| | White | 7.98 (1.40) | 7.91 (1.59) | 4.66 (0.93) | 6.09 (0.78) | 5.30 (0.90) | 5.86 (1.19) | 7.72 (1.75) | 9.32 (2.46) | 4.95 (1.24) |
| Bronx | Black | 7.47 (2.09) | 12.48 (2.35) | 8.94 (2.94) | 9.08 (1.42) | 6.53 (1.82) | 7.26 (1.72) | 7.08 (2.54) | 7.29 (1.83) | 11.43 (3.48) |

| | | | | | | | | | | |
|---|---|---|---|---|---|---|---|---|---|---|
| | Latina | 10.82 (2.08) | 14.78 (2.96) | 14.41 (1.75) | 15.21 (1.82) | 12.79 (1.99) | 12.39 (2.30) | 15.04 (2.35) | 13.21 (1.95) | 13.83 (1.74) |
| | White | 12.66 (8.28) | 15.29 (7.59) | 12.81 (7.90) | 4.74 (4.77) | 9.35 (5.10) | 3.42 (2.80) | 5.53 (6.35) | 4.57 (2.10) | 9.50 (5.20) |
| Brooklyn | Black | 11.51 (2.25) | 10.35 (1.38) | 8.96 (1.17) | 10.67 (1.34) | 12.01 (2.05) | 7.26 (3.24) | 11.66 (1.75) | 11.79 (2.35) | 12.09 (2.86) |
| | Latina | 9.61 (1.98) | 7.49 (2.79) | 11.95 (2.04) | 20.75 (3.05) | 11.65 (1.02) | 16.57 (5.17) | 10.55 (1.80) | 13.72 (1.65) | 8.78 (2.42) |
| | White | 4.03 (2.12) | 6.12 (2.71) | 4.07 (1.73) | 4.90 (0.63) | 7.20 (2.43) | 5.53 (1.85) | 1.31 (0.77) | 7.22 (2.29) | 8.41 (2.62) |
| Manhattan | Black | 9.30 (2.08) | 16.14 (3.91) | 7.73 (2.81) | 7.93 (2.57) | 10.17 (2.56) | 5.21 (1.34) | 14.48 (6.46) | 10.69 (1.45) | 8.65 (1.99) |
| | Latina | 18.40 (2.09) | 18.00 (3.50) | 14.61 (2.62) | 11.86 (2.72) | 13.34 (1.67) | 13.23 (1.33) | 11.50 (1.58) | 10.37 (1.10) | 8.85 (1.42) |
| | White | 10.52 (4.25) | 4.73 (5.18) | 5.59 (2.04) | 6.06 (2.85) | 2.76 (1.79) | 6.07 (4.19) | 15.64 (3.04) | 13.20 (4.81) | 3.42 (1.40) |
| Queens | Black | 7.42 (2.52) | 8.14 (2.14) | 1.79 (1.25) | 12.65 (2.06) | 12.34 (2.88) | 9.08 (2.54) | 4.24 (1.53) | 13.13 (3.13) | 6.71 (2.35) |
| | Latina | 17.22 (2.85) | 12.64 (2.10) | 15.20 (1.52) | 11.87 (2.36) | 7.25 (1.25) | 12.66 (1.34) | 13.16 (2.53) | 16.35 (1.61) | 8.55 (1.23) |
| | White | 16.55 (3.28) | 9.04 (3.91) | 1.45 (1.39) | 7.26 (1.52) | 4.58 (1.81) | 6.73 (2.69) | 8.81 (3.29) | 16.18 (9.72) | 0.00 (0.00) |
| Staten Island | Black | 8.49 (3.56) | 12.10 (3.92) | 2.99 (1.67) | 14.49 (6.39) | 6.24 (2.86) | 5.94 (2.14) | 10.86 (4.02) | 8.77 (4.42) | 9.19 (3.37) |
| | Latina | 14.39 (3.68) | 13.04 (3.81) | 12.77 (2.19) | 16.19 (3.70) | 8.83 (1.79) | 13.60 (2.09) | 16.28 (2.57) | 8.93 (2.38) | 13.42 (2.97) |
| | White | 8.07 (1.32) | 8.35 (1.72) | 5.73 (1.15) | 7.00 (1.38) | 5.37 (1.37) | 5.67 (1.44) | 7.17 (2.01) | 3.57 (1.26) | 5.18 (1.67) |

**Supplementary Table 5**. Suicide Attempt Odds Ratios by NYC Borough and Race/Ethnicity Among Female Adolescents, 2003–2019

| | | **Black/African American** | | | **Hispanic/Latina** | | | **White** | | |
|---|---|---|---|---|---|---|---|---|---|---|
| | | **OR** | **95% CI** | ***P*** | **OR** | **95% CI** | ***P*** | **OR** | **95% CI** | ***P*** |
| **Bronx** | **L** | 0.93 | 0.47 - 1.85 | 0.83 | 1.06 | 0.72 - 1.54 | 0.78 | 0.38 | 0.08 - 1.91 | 0.23 |
| | **Q** | 1.30 | 0.74 - 2.29 | 0.36 | 0.88 | 0.63 - 1.23 | 0.46 | 1.60 | 0.31 - 8.21 | 0.56 |
| | **C** | 1.71 | 0.91 - 3.22 | 0.10 | 1.22 | 0.81 - 1.84 | 0.33 | 2.05 | 0.49 - 8.59 | 0.31 |
| **Brooklyn** | **L** | 1.10 | 0.73 - 1.66 | 0.64 | 1.15 | 0.66 - 2.01 | 0.62 | 1.66 | 0.69 - 3.97 | 0.25 |
| | **Q** | 1.24 | 0.77 - 2.00 | 0.37 | **0.49** | **0.29 - 0.83** | **0.01** | 1.25 | 0.53 - 2.93 | 0.60 |

| | | | | | | | | | | |
|---|---|---|---|---|---|---|---|---|---|---|
| | **C** | 0.96 | 0.60 - 1.51 | 0.85 | 0.96 | 0.56 - 1.67 | 0.90 | 1.57 | 0.67 - 3.69 | 0.29 |
| **Manhattan** | **L** | 0.89 | 0.42 - 1.90 | 0.77 | **0.47** | **0.32 - 0.70** | **<0.001** | 1.25 | 0.47 - 3.34 | 0.65 |
| | **Q** | 1.20 | 0.72 - 2.00 | 0.47 | 1.03 | 0.74 - 1.43 | 0.85 | 1.02 | 0.28 - 3.68 | 0.97 |
| | **C** | 0.92 | 0.41 - 2.06 | 0.84 | 0.88 | 0.62 - 1.25 | 0.48 | **0.22** | **0.07 - 0.67** | **0.01** |
| **Queens** | **L** | 1.23 | 0.64 - 2.39 | 0.53 | 0.76 | 0.52 - 1.12 | 0.16 | 0.70 | 0.19 - 2.63 | 0.59 |
| | **Q** | 0.78 | 0.38 - 1.57 | 0.47 | 1.21 | 0.84 - 1.74 | 0.29 | 1.64 | 0.81 - 3.34 | 0.16 |
| | **C** | 0.78 | 0.40 - 1.55 | 0.47 | 0.69 | 0.47 - 1.01 | 0.06 | **0.19** | **0.07 - 0.51** | **0.002** |
| **Staten Island** | **L** | 1.07 | 0.41 - 2.82 | 0.89 | 0.91 | 0.52 - 1.62 | 0.76 | **0.57** | **0.35 - 0.93** | **0.02** |
| | **Q** | 1.31 | 0.55 - 3.12 | 0.55 | 1.03 | 0.62 - 1.72 | 0.90 | 1.05 | 0.63 - 1.75 | 0.84 |
| | **C** | 0.90 | 0.37 - 2.18 | 0.82 | 0.96 | 0.58 - 1.60 | 0.89 | 0.94 | 0.53 - 1.68 | 0.84 |
